# Association between fermented milk consumption and oesophageal carcinoma among patients presenting to two referral hospitals in Nandi County, Kenya: a case-control study

**DOI:** 10.1101/2025.04.26.25326497

**Authors:** Rancy C. Mutai, Marshal M. Mweu

## Abstract

**Background:** Oesophageal carcinoma (OC) is a prominent cause of morbidity and mortality in low and middle-income countries yet little evidence exists about its contextual drivers. The objectives of this study were to assess the association between fermented milk (FM) (*mursik*) consumption and OC independent of known socio-demographic and lifestyle risk factors as well as to quantify its population impact among patients presenting to two referral hospitals in Nandi County, Kenya.

**Methods:** A hospital-based case-control study was employed to assess the FM-OC relationship among patients presenting to Kapsabet Surgical Care Centre and Kapsabet County Referral Hospital in Nandi County, Kenya for care between 23^rd^ November 2023 and 13^th^ January 2024. All 33 cases meeting specific eligibility criteria were prospectively recruited whilst 131 controls were simple randomly sampled and frequency-matched to the cases by hospital and day of presentation. A logistic regression model was fitted to assess the FM-OC association while adjusting for potential confounders. Subsequently, a population attributable fraction (PAF) for the association (along with its confidence interval) was estimated.

**Results:** A strong association between FM and OC was noted; the odds of OC among frequent and infrequent consumers of FM being over nine (OR 9.1, 95% CI: 3.1-26.6) and three (OR 3.2, 95% CI: 1.1-9.2) times higher, respectively, than non-consumers. This association was not substantially confounded by the studied socio-demographic and lifestyle factors. The PAF estimate for this association was 65.2% (95% CI: 40.1-90.7).

**Conclusions:** In this study setting, FM consumption was strongly associated with OC independent of other risk factors. This association registered a high PAF suggesting that up to 65% of OC in the population could be prevented if FM was not consumed. This finding calls for safe, community-owned alternatives for fermenting milk in order to mitigate the risk of OC in this population.

## Introduction

Gastrointestinal (GI) cancers constitute a major public health concern accounting for 26% of the global cancer incidence and 35% of all cancer-related deaths in 2018[1]. Of GI cancers, oesophageal carcinoma (OC) features prominently particularly in low and middle-income countries (LMICs) [2]. OC is a malignancy of the oesophagus typically characterised by dysphagia, odynophagia, unexplained weight loss, chest pain, worsening dyspepsia, and chronic cough[3].

In Sub-Saharan Africa, the highest incidence of OC is registered in the ‘oesophageal cancer corridor’ – an area stretching from Ethiopia to South Africa and traversing Kenya, Uganda, Tanzania and Malawi [4]. In Eastern Africa, OC incidence averages about 8.3 per 100,000 person-years [1]– Kenya recording an age-standardised incidence of 28.7 cases per 100,000 [5] and a case fatality rate of 99%[6].

Fermented milk (FM) (commonly known as *mursik*) remains a favourite cultural beverage among the indigenous inhabitants of the Rift Valley region of Kenya [7,8]. It is prepared by boiling and fermenting fresh cow’s or goat’s milk for a period of 3-5 days, and thereafter, charcoal powder is added[7,8]. Despite the health benefits of fermented milk, its consumption has been linked to the occurrence of OC [9] ascribable to improper milk handling and fermentation processes which may introduce contaminants such as moulds that produce various mycotoxins[7,10]. These toxins may contribute to oesophageal carcinogenesis when ingested habitually[8,10]. Moreover, charcoal as a key ingredient is believed to harbour polycyclic aromatic hydrocarbons (PAHs)[11] that could catalyse the carcinogenicity of *mursik*.

Besides FM, alcohol and tobacco use, biomass fuels, socio-demographic characteristics and hot beverage consumption are well-recognised risk factors for OC [9,12–17]. Alcoholic drinks may contain acetaldehydes and nitrosamines that are carcinogenic [9,18]. PAHs, aldehydes, and nitrosamines released during tobacco smoking may precipitate carcinogenesis[19]. In Kenya, wood fuel, ubiquitously used in rural households, is known to emit PAHs, benzene, arsenic, 1,3-butadiene and formaldehyde that can trigger oesophageal carcinogenesis[16,17,20,21]. The risk of OC increases with age, with individuals aged ≥50 years bearing a disproportionate risk [13]. OC is more common in males than females – attributable to higher rates of tobacco smoking and alcohol use among males[22,23]. Consumption of hot beverages >65^0^c could potentially be carcinogenic[24]. Hot drinks may instigate thermal injury of the oesophageal mucosa – a risk for OC[25,26].

Despite OC’s preponderance in LMICs, there is scarcity of evidence on its contextual drivers in endemic settings. Consequently, the objectives of this study were to quantify the association between FM consumption and OC independent of known risk factors as well as to estimate FM’s population impact among patients presenting to two referral hospitals in Nandi County, Kenya. An understanding of FM’s role in the occurrence of OC is critical to guiding policy development and informing the design of appropriate interventions for the control of OC in Kenya.

## Materials and Methods

### Study setting, design and population

The study was carried out at Kapsabet Surgical Care Centre (KSCC) and Kapsabet County Referral Hospital (KCRH) situated in Kapsabet town, Nandi County within the Rift Valley region of Kenya. KSCC is a private hospital specialising in endoscopy and surgical care whereas KCRH is a state-run referral facility offering a range of preventive and curative services. Notably, the two hospitals serve a catchment population of ∼ 885,700 persons comprising majorly of Nandi natives of the Kalenjin community [27]. Of note, subsistence and commercial crop (tea and maize) and dairy farming are the economic mainstay of this region.

A hospital-based case-control study design was employed in this study to evaluate the association between FM consumption and OC. The rationale for the choice of the design stems not only from its suitability for studying rare outcomes but also the ease and efficiency of recruitment of cases presenting to a hospital setting for care. Importantly, the study conformed to STROBE guidelines for reporting of case-control studies [28].

The study population comprised County-resident patients aged ≥18 years presenting to KSCC and KCRH for care between 23^rd^ November 2023 and 13^th^ January 2024 and who had consented to participate in the study. Critically-ill patients and those uninterviewable and/or unable to give consent were excluded from the study.

### Outcome definition

Cases were eligible individuals attending the facilities with any suggestive symptoms: dysphagia, odynophagia, persistent coughing, chest pains and unexplained weight loss and had a confirmed diagnosis of OC during the study period. Controls were patients presenting to the same facilities as cases but with other conditions.

### Sample size and participant selection

The size of the study sample was estimated as suggested by Kelsey *et al*. [29] for case control studies:

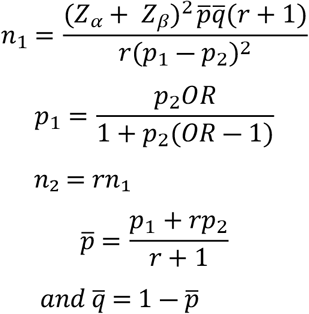

Where *Z*_*α*_ = 1.96 is the required value for a two-tailed 95% confidence level; *Z*_*β*_ = -0.84 is the desired value for an 80% statistical power; *n*_1_ is the number of cases; *n*_2_ is the number of controls; *r* = 4 is the ratio of controls to cases; *p*_1_ is the proportion of cases consuming FM; *p*_2_ is the proportion of controls that consume FM – specified at 10.5% based on a previous study[9] and *OR* is the odds ratio for the FM-OC association (set at 3.72) informed by literature[9]. Given an anticipated non-response rate of 5%, the necessary sample was 164 subjects: 33 cases and 131 controls. All eligible cases were prospectively recruited over the study period within the facilities until the desired number was attained. Controls were simple randomly sampled from a sampling frame of patients admitted at the facilities’ medical and surgical wards and frequency-matched to cases by hospital and day of presentation.

### Study variables

The primary exposure in this study was FM consumption. The other exposures considered included the participants’ sociodemographic characteristics (age, sex, education level, employment status and marital status), lifestyle factors (alcohol, tobacco use and beverage temperature) and cooking fuel. These variables were measured using an orally administered semi-structured questionnaire captured in both English and Swahili. Table 1 displays the assessment of these variables.

**Table 1.** Measurement of the study variables.

| Variable (type) | Measurement |
| --- | --- |
| Fermented milk (ordinal) | The respondent's consumption frequency of fermented milk was captured in three categories: non-consumer, infrequent (1-2 times a week) and frequent ($\geq 3$ times a week) |
| Alcohol (ordinal) | The respondent's consumption frequency of alcohol was classified into three categories: non-consumer, infrequent (1-2 times a week) and frequent ($\geq 3$ times a week) |
| Cooking fuel (nominal) | This reflected the respondent's primary type of cooking fuel. Categorised into three: biomass fuels, natural/biogas and kerosene |
| Tobacco (nominal) | Assessed on the basis of smoking or chewing of tobacco: user or non-user. |
| Beverage temperature (ordinal) | This was premised on the respondent's preferred consumption temperature of beverages. Classified into three categories: warm, hot and very hot. |
| Age (continuous) | Captured in years. |
| Sex (nominal) | Captured as male or female. |
| Marital status (nominal) | Assessed in four categories: single, married, divorced/separated or widowed |
| Employment status (nominal) | Captured in two categories: employed or unemployed. |
| Education level (ordinal) | This reflected an individual's highest level of education: no formal education, primary, secondary and tertiary levels. |

### Ethical considerations

Written permission to conduct the study was secured from the Kenyatta National Hospital-University of Nairobi Ethics and Research Committee (PF26/06/2023), the National Commission for Science, Technology and Innovation (NACOSTI/P/23/30502) and the Nandi County Health Services (Ref. No. CDH/NDI/2021/R.A/5). Furthermore, written informed consent was obtained from the participants prior to enlisting them in the study. In addition, the data collected were analyzed anonymously.

### Minimisation of biases

Before data collection, two research assistants were trained on standardised interviewing techniques in order to limit interviewer bias during elicitation of information from respondents. Since differential recall of consumption habits (FM, alcohol and beverage) between cases and controls was likely, questioning on these related to recent consumption. Moreover, as the likelihood of altering consumption patterns by cases was probable, reverse causality was minimised by focusing on incident cases.

### Statistical analysis

Upon data collection, the questionnaire responses were examined for completeness, after which qualitative variables were coded. Subsequently, the data were entered into a Microsoft Excel spreadsheet and cross-checked for accuracy. The dataset was then exported to R software v4.4.0 for cleaning and analysis. The R code for these analyses is available as supporting information[30]. For descriptive statistics, medians and ranges were computed for continuous variables whereas frequencies and percentages summarised categorical variables. A logistic regression model was fitted to evaluate the crude association between FM and OC. To evaluate the study covariates (age, sex, education level, employment status, marital status, alcohol, cooking fuel, tobacco and beverage temperature) as potential confounders of the FM–OC association, each of these factors was separately screened for its association with OC and FM using the Fisher’s exact test at *P*<0.05 (this two-stage testing process is synonymous with sequential screening where only variables associated with the outcome at *P*<0.05 are subjected to subsequent assessment with the primary exposure). For analytical ease, age was grouped into three categories: ≤40 years, 41 – 65 years, >65 years and cooking fuel was reclassified into two categories: biomass/kerosene and natural/biogas since there were no cases using kerosene. Qualifying covariates from this sequential screening were deemed potential confounders of the FM–OC relationship and thus included in a multivariable logistic regression model to adjust for their confounding effect. At this stage, only those variables that resulted in a >20% change in the coefficient for FM following a backward step-wise elimination procedure were considered important confounders and thus retained in the final model [31].

A population attributable fraction (PAF) – a measure quantifying the proportion of disease in the population that is preventable if the exposure were eliminated[31]– was computed from estimates of the final model [32]:

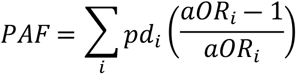

Where: *pd*_*i*_ reflects the proportion of cases consuming FM infrequently and frequently (non-baseline categories) and *a OR* _*i*_ is the specific adjusted odds ratio for the non-baseline levels of FM. Estimation of PAF (along with its bootstrapped confidence interval) was realised using the *graphPAF* package [33]written in R software.

## Results

Of the sample of 164 subjects, 160 (32 cases and 128 controls) gave consent to the study. Descriptive statistics for the study exposure variables are indicated in Table 2. The respondents’ median age was 60 years (Range: 30-78) for cases and 50 years (Range: 21-98) for controls. On FM, 46.9% (*n* = 15) of cases were frequent consumers compared with 14.8% (*n* = 19) of controls. About 28.1% (*n* = 9) of cases were frequent consumers of alcohol in comparison with 10.2% (*n* = 13) of controls.

**Table 2.** Summary statistics for the socio-demographic and lifestyle characteristics of the respondents.

| Variable | Category | Cases: *n* (%) | Controls: *n* (%) |
| --- | --- | --- | --- |
| Age in years | Median (Range)* | 60 (30 - 78) | 50 (21 - 98) |
| Sex | Male | 21 (65.6) | 60 (46.9) |
|  | Female | 11 (34.4) | 68 (53.1) |
| Marital status | Single | 2 (6.3) | 29 (22.7) |
|  | Married | 24 (75.0) | 85 (66.4) |
|  | Divorced/Separated | 4 (12.5) | 6 (4.7) |
|  | Widowed | 2 (6.3) | 8 (6.3) |
| Education level | No formal education | 4 (12.5) | 11 (8.6) |
|  | Primary | 21 (65.6) | 48 (37.5) |
|  | Secondary | 6 (18.8) | 35 (27.3) |
|  | Tertiary | 1 (3.1) | 34 (26.6) |
| Employment status | Unemployed | 15 (46.9) | 56 (43.8) |
|  | Employed | 17 (53.1) | 72 (56.3) |
| Alcohol | Non-consumer | 10 (31.2) | 85 (66.4) |
|  | Infrequent | 13 (40.6) | 30 (23.4) |
|  | Regular | 9 (28.1) | 13 (10.2) |
| Tobacco | Non-user | 17 (53.1) | 109 (85.2) |
|  | User | 15 (46.9) | 19 (14.8) |
| Beverage temperature | Warm | 18 (56.2) | 66 (51.6) |
|  | Hot | 7 (21.9) | 41 (32.0) |
|  | Very hot | 7 (21.9) | 21 (16.4) |
| Fermented milk | Non-consumer | 6 (18.8) | 69 (53.9) |
|  | Infrequent | 11 (34.4) | 40 (31.3) |
|  | Frequent | 15 (46.9) | 19 (14.8) |
| Cooking fuel | Biomass | 27 (84.4) | 83 (64.8) |
|  | Kerosene | 0 (0.0) | 8 (6.3) |
|  | Natural/biogas | 5 (15.6) | 37 (28.9) |

\*Median (Range): only applies to age

The crude association between FM and OC is captured in Table 3. The odds of OC were approximately three (3.2; 95% CI: [1.1-9.2]) and nine (9.1; 95% CI: [3.1-26.6]) times higher amongst infrequent and frequent consumers, respectively, than non-consumers.

**Table 3.** Univariable analysis of the association between fermented milk consumption and oesophageal carcinoma among patients at KSCC and KCRH, Nandi County, Kenya.

| Variable | Category | cOR* | 95% CI | P-value |
| --- | --- | --- | --- | --- |
| Fermented milk | Non-consumer | Ref | - | <0.001 |
|  | Infrequent | 3.2 | 1.1; 9.2 |  |
|  | Frequent | 9.1 | 3.1; 26.6 |  |

\*Crude odds ratio

The association between individual covariates and OC is presented in Table 4. Notably, age group, education level, alcohol and tobacco were associated with OC at the 5% significance level. As such, these variables qualified for assessment of their association with FM.

**Table 4.** Association between individual covariates and oesophageal carcinoma among patients attending KSCC and KCRH, Nandi County, Kenya.

| Variable | Category | Cases: *n* (%) | Controls: *n* (%) | P-value |
| --- | --- | --- | --- | --- |
| *Age group | ≤ 40 years | 2 (6.3) | 42 (32.8) | 0.005 |
|  | 41-65 years | 21 (65.6) | 57 (44.5) |  |
|  | >65 years | 9 (28.1) | 29 (22.7) |  |
| Sex | Male | 21 (65.6) | 60 (46.9) | 0.075 |
|  | Female | 11 (34.4) | 68 (53.1) |  |
| *Education level | No formal education | 4 (12.5) | 11 (8.6) | 0.003 |
|  | Primary | 21 (65.6) | 48 (37.5) |  |
|  | Secondary | 6 (18.8) | 35 (27.3) |  |
|  | Tertiary | 1 (3.1) | 34 (26.6) |  |
| Employment status | Unemployed | 15 (46.9) | 56 (43.8) | 0.843 |
|  | Employed | 17 (53.1) | 72 (56.2) |  |
| Marital status | Single | 2 (6.3) | 29 (22.7) | 0.066 |
|  | Married | 24 (75.0) | 85 (66.4) |  |
|  | Divorced/separated | 4 (12.5) | 6 (4.7) |  |
|  | Widowed | 2 (6.3) | 8 (6.3) |  |
| *Alcohol | Non-consumers | 10 (31.3) | 85 (66.4) | 0.001 |
|  | Infrequent | 13 (40.6) | 30 (23.4) |  |
|  | Frequent | 9 (28.1) | 13 (10.2) |  |
| Cooking fuel | Biomass/kerosene | 27 (84.4) | 91 (71.1) | 0.177 |
|  | Natural/biogas | 5 (15.6) | 37 (28.9) |  |
| *Tobacco | Non-users | 17 (53.1) | 109 (85.2) | <0.001 |
|  | users | 15 (46.9) | 19 (14.8) |  |
| Beverage temperature | Warm | 18 (56.3) | 66 (51.6) | 0.480 |
|  | Hot | 7 (21.9) | 41 (32.0) |  |
|  | Very hot | 7 (21.9) | 21 (16.4) |  |

\*Eligible for screening for their association with fermented milk ( $p < 0.05$ )

Table 5 presents the results for the association between qualifying covariates and FM. Importantly, only age group and alcohol were significantly associated with fermented milk (*p* <0.05). Consequently, these variables were deemed potential confounders of the FM-OC association and were offered to a multivariable logistic regression model to adjust for their confounding effect.

**Table 5.** Association between qualifying exposures and fermented milk consumption among patients attending KSCC and KCRH, Nandi County, Kenya.

| Variable | Category | Non-consumers:<br>*n* (%) | Infrequent consumers: *n* (%) | Frequent consumers: *n* (%) | *P*-value |
| --- | --- | --- | --- | --- | --- |
| *Age group | ≤ 40 years | 26(34.7) | 12(23.5) | 6(17.6) | 0.038 |
|  | 41-65 years | 30(40.0) | 32(62.7) | 16(47.1) |  |
|  | >65 years | 19(25.3) | 7(13.7) | 12(35.3) |  |
| Education level | No formal education | 6(8.0) | 4(7.8) | 5(14.7) | 0.096 |
|  | Primary | 27(36.0) | 21(41.2) | 21(61.8) |  |
|  | Secondary | 22(29.3) | 14(27.5) | 5(14.7) |  |
|  | Tertiary | 20(26.7) | 12(23.5) | 3(8.8) |  |
| *Alcohol | Non-consumers | 54(72.0) | 31(60.8) | 10(29.4) | 0.001 |
|  | Infrequent | 15(20.0) | 13(25.5) | 15(44.1) |  |
|  | Frequent | 6(8.0) | 7(13.7) | 9(26.5) |  |
| Tobacco | Non-users | 63(84.0) | 39(76.5) | 24(70.6) | 0.250 |
|  | users | 12(16.0) | 12(23.5) | 10(29.4) |  |

\*Eligible for inclusion in the multivariable logistic regression model ( $p < 0.05$ )

Results of the multivariable analysis of the FM-OC association are presented in Table 6. Since alcohol and age group did not confound the FM-OC association (i.e. neither of the factors resulted in a >20% change in the coefficient for FM) the odds ratio for the unadjusted association was maintained and subsequently employed in the derivation of PAF. The PAF estimate for the FM-OC association was 65.2% (95% CI: 40.1-90.7).

**Table 6:** Multivariable analysis of the association between fermented milk and oesophageal carcinoma among patients at KSCC and KCRH, Nandi County, Kenya.

| Variable | Category | aOR* | 95% CI | P-value |
| --- | --- | --- | --- | --- |
| Fermented milk | Non-consumer | Ref | - | 0.005 |
|  | Infrequent | 2.5 | 0.8; 7.7 |  |
|  | Frequent | 6.1 | 2.0; 19.2 |  |
| Alcohol | Non-consumer | Ref | - | 0.085 |
|  | Infrequent | 2.4 | 0.9; 6.6 |  |
|  | Frequent | 3.3 | 1.0; 10.8 |  |
| Age group | ≤ 40 years | Ref | - | 0.026 |
|  | 41-65 years | 6.3 | 1.3; 30.3 |  |
|  | >65 years | 4.2 | 0.8; 22.9 |  |

\*Adjusted odds ratio

## Discussion

In this population, an analysis of the association between FM consumption and OC has demonstrated a robust relationship. The odds of OC were roughly three and nine times higher amongst infrequent and frequent consumers, respectively, than non-consumers; signifying a dose-response relationship. In particular, this association was not confounded by any of the examined covariates. Patel et al. [2013][9] corroborate this strong relationship – observing about four times higher odds of OC amongst *mursik* consumers. Importantly, markedly high concentrations of ethanol (>100mM) and acetaldehyde (>1800µM) have been detected previously in *mursik* [10] – carcinogenicity of acetaldehyde being exhibited in both animal and in vitro models at fairly low concentrations (100 µM) [34,35]Conceivably, recurrent exposure of the oesophageal mucosa to these mutagenic substances in *mursik* may explain its carcinogenicity. In Afghanistan, a linkage between consumption of fermented dairy products (cheese and yoghurt) and OC has been established[36].

As sterility during the traditional fermentation process is not guaranteed, contamination of *mursik* by fungal species of the genera *Aspergillus, Penicillium* and *Fusarium* is likely, with resultant production of mycotoxins [7]. Of these, aflatoxin is the predominant metabolite recovered from *mursik*[37]. Notably, aflatoxins have been implicated in oesophageal carcinogenesis [38].

Customarily, fermentation gourds for *mursik* are lined with specially ground fine charcoal that imparts a smoky flavour[39]. Nonetheless, charcoal could be a source of PAHs – potentially carcinogenic – spurring oesophageal carcinogenesis[11]. More so, charcoal-derived carcinogens have been pointed out as contributory to the high incidence of OC in Iran [40].

Despite our finding of a significant elevated risk of OC with FM consumption, some studies have posted contradictory findings[19,41]. In particular, Oncina-Cánovas et al. [41] report a protective association between consumption of fermented dairy products and OC. This divergence of observations across study contexts underscores the need to carefully examine the role played by disparate dairy fermentation processes in driving local and/or regional OC burdens.

The PAF estimate of 65.2% for FM in this study implies that roughly two-thirds of OC in the study population could potentially be prevented if FM was not consumed. Irrefutably, FM as a singular factor is impactful in this population – contributing significantly to the OC burden in this setting. This highlights the necessity of raising awareness on general safety practices for preparing fermented milk.

A few limitations are inherent in this study. Despite efforts to adjust the association for known confounders, the possibility of residual confounding persists. Other unmeasurable factors such as genetic predispositions and environmental exposures could still have influenced the observed relationship between FM and OC. Although the recruitment of controls from hospital settings not only affords convenience but also controls for potential differences in health-seeking behaviours between cases and controls, their utility may introduce selection bias. Hospitalised controls generally tend to have higher exposures compared to the general population that they represent. Consequently, for our study, this undertaking is likely to have weakened the FM-OC association.

## Conclusions

This study has revealed a strong association between FM consumption and OC in this setting. This relationship was not confounded by any of the studied factors. The association yielded a high PAF implying that up to 65% of OC in the population could be prevented if FM was not consumed. This finding calls for dedicated campaigns to raise local awareness with a view to develop safe, community-acceptable fermentation processes to mitigate OC risk.

## Data Availability

All relevant data are within the manuscript and its Supporting Information files.

https://dataverse.harvard.edu/dataset.xhtml?persistentId=doi:10.7910/DVN/DC0IMK

## Acknowledgments

The authors heartily thank the study participants, the administration and staff at KSCC and KCRH for making it possible to accomplish this research work.

